# A computational biomarker of juvenile myoclonic epilepsy from resting-state MEG

**DOI:** 10.1101/2020.05.18.20102681

**Authors:** Marinho A. Lopes, Dominik Krzemiński, Khalid Hamandi, Krish D. Singh, Naoki Masuda, John R. Terry, Jiaxiang Zhang

**Affiliations:** Cardiff University Brain Research Imaging Centre, School of Psychology, Cardiff University, Cardiff CF24 4HQ, United Kingdom; The Welsh Epilepsy Unit, Department of Neurology, University Hospital of Wales, Cardiff CF14 4XW, United Kingdom; Department of Mathematics, University at Buffalo, State University of New York, USA; Computational and Data-Enabled Science and Engineering Program, University at Buffalo, State University of New York, USA; EPSRC Centre for Predictive Modelling in Healthcare, University of Birmingham, Birmingham, United Kingdom; Centre for Systems Modelling and Quantitative Biomedicine, University of Birmingham, Edgbaston, United Kingdom; Institute for Metabolism and Systems Research, University of Birmingham, Edgbaston, United Kingdom

**Keywords:** epilepsy diagnosis, juvenile myoclonic epilepsy, biomarker, MEG, functional connectivity, phenomenological model

## Abstract

**Objective:** Functional networks derived from resting-state scalp EEG from people with idiopathic (genetic) generalized epilepsy (IGE) have been shown to have an inherent higher propensity to generate seizures than those from healthy controls when assessed using the concept of brain network ictogenicity (BNI). Herein we test whether the BNI framework is applicable to resting-state MEG and whether it may achieve higher classification accuracy relative to previous studies using EEG.

**Methods:** The BNI framework consists in deriving a functional network from apparently normal brain activity, placing a mathematical model of ictogenicity into the network and then computing how often such network generates seizures in silico. We consider data from 26 people with juvenile myoclonic epilepsy (JME) and 26 healthy controls.

**Results:** We find that resting-state MEG functional networks from people with JME are characterized by a higher propensity to generate seizures (i.e. BNI) than those from healthy controls. We found a classification accuracy of 73%.

**Conclusions:** The BNI framework is applicable to MEG and capable of differentiating people with epilepsy from healthy controls. The observed classification accuracy is similar to previously achieved in scalp EEG.

**Significance:** The BNI framework may be applied to resting-state MEG to aid in epilepsy diagnosis.

**Highlights:**

- Computational modelling is combined with MEG to differentiate people with juvenile myoclonic epilepsy from healthy controls.
- Brain network ictogenicity (BNI) was found higher in people with juvenile myoclonic epilepsy relative to healthy controls.
- BNI’s classification accuracy was 73%, similar to previously observed using scalp EEG.

## 1. Introduction

Epilepsy is one of the most common neurological disorders with an estimated 5 million new diagnosis each year (WHO, 2019). The diagnosis of epilepsy is based on clinical history and supported by clinical electroencephalography (EEG). The presence of interictal spikes in the routine scalp EEG recordings is one of the most valuable biomarkers of epilepsy (Pillai and Sperling, 2006). However, the presence of interictal epileptiform discharges (IED) in a routine EEG is low, ranging between 25 and 56% (Smith, 2005; Benbadis et al., 2020). Furthermore, about 10% of people with epilepsy do not show IEDs even after repeated or prolonged EEG (Smith, 2005;Benbadis et al., 2020). On the other hand, specificity is also suboptimal, ranging between 78 and 98% (Smith, 2005), which, for example, may delay the diagnosis of psychogenic nonepileptic attacks by 7 to 10 years (Benbadis, 2009).

The low sensitivity of IEDs results from IEDs being typically rare events. This may be a consequence of their sources being deep in the brain and/or the extent of cortex involved in epileptic activity being undetectable at the scalp surface (Pillai and Sperling, 2006). Consequently, much of the routine clinical EEG recording consists of brain activity that appears normal to visual inspection, which without other visible disturbances in background rhythms is considered non-informative. However, growing evidence suggests that such sections of interictal EEG without IEDs may be used to inform epilepsy diagnosis (e.g. Larsson and Kostov, 2005; Schmidt et al., 2016; Verhoeven et al., 2008).Larsson and Kostov (2005) showed that there is a shift in the peak of the alpha power towards lower frequencies in interictal EEG from people with both focal and generalized epilepsy. More recently, Abela et al. (2019) found that a slower alpha rhythm may be an indicator of seizure liability. Other studies have used graph theory to test whether functional networks derived from interictal EEG differ from EEG obtained from healthy controls. It has been found that functional networks from people with epilepsy are more “regular” (i.e. higher path lengths between nodes) and deviate more from small-world structures than those found in healthy controls (Horstmann et al., 2010; Quraan et al., 2013).Functional network alterations inferred from resting-state EEG have also been used to differentiate children with focal epilepsy from healthy children (van Diessen et al., 2013,2016). Furthermore, resting-state EEG functional networks from people with IGE were shown to have more functional connections than healthy controls (Chowdhury et al., 2014). Functional networks inferred from interictal EEG from people with temporal lobe epilepsy have also been shown to differ from those from healthy controls (Coito et al., 2016).

All these studies show that functional networks based on apparently normal EEG may aid in the diagnosis of epilepsy. However, these studies lack mechanistic insights as to why such differences may be related to epilepsy. To build such understanding, we and others have proposed to use mathematical models of epilepsy to assess the functional networks and elucidate as to why a brain may be prone to generate seizures (Schmidt et al., 2014, 2016; Petkov et al., 2014; Lopes et al., 2019). In particular, we showed that resting-state EEG functional networks from people with IGE are more prone to support synchronization phenomena and the emergence of seizure-like activity than those from controls (Schmidt et al., 2014; Petkov et al., 2014). To quantify the differences, we introduced the concept of brain network ictogenicity (BNI), i.e. a measure of how likely a functional network is of generating seizures *in silico* (Petkov et al., 2014).

For the BNI to be useful for diagnosing people with epilepsy from apparently normal brain activity, we relied on the assumption that the ability of a brain to generate seizures is an enduring feature that should be identifiable during interictal periods. We further assumed that such underlying closeness to seizures is captured by the properties of functional networks. We then assess the capacity of a given functional network to generate seizures by estimating BNI through computer simulations that produce long-term activity from which the volume of epileptiform activity can be evaluated. People with epilepsy were therefore assumed to have resting-state functional networks that were more ictogenic, i.e. that had a higher propensity to generate seizures as estimated by the BNI, compared to healthy people. Using this framework on a dataset comprising 30 people with IGE and 38 healthy controls it was found 100% specificity at 56.7% sensitivity, and 100% sensitivity at 65.8% specificity (Schmidt et al., 2016).

In the current study, we aim to test whether the BNI concept may be equally useful when applied to resting-state MEG data (i.e. its generalizability to a different data modality), and whether it may yield superior diagnostic power of epilepsy relative to previous applications of BNI to resting-state EEG data. In particular, we aim to find whether BNI may be capable of differentiating juvenile myoclonic epilepsy (JME) from healthy controls, using MEG data, and observe how classification accuracy compares to previous studies of BNI on scalp EEG (Schmidt et al., 2014, 2016). Since MEG has the advantage, relative to EEG, of neuromagnetic fields being minimally perturbed by brain tissue, skull and scalp (Supek and Aine, 2016), one may expect that MEG-derived functional networks may be more reliable than those from EEG, which in turn may enhance the BNI’s ability to differentiate people with generalized epilepsy from healthy controls.

## 2. Methods

### 2.1 Participants

We used resting-state MEG data obtained from 26 people with JME and 26 healthy controls. The individuals with epilepsy were recruited from a specialist clinic for epilepsy at University Hospital of Wales in Cardiff, and the healthy individuals were volunteers who had no history of significant neurological or psychiatric disorders. The healthy group was age and gender matched to the epilepsy group. The age range in the epilepsy group was 17 to 47, median 27 years, and in the control group was 18 to 48, median 27 years. There were 7 males in the epilepsy group and 7 males in the control group. Individuals in the epilepsy group had a number of different seizure types and were taking anti-epileptic drugs (see Krzemiński et al. (2020) and Routley et al. (2020) for more details about this dataset). Table 1 summarizes the clinical characteristics of the individuals with epilepsy. This study was approved by the South East Wales NHS ethics committee, Cardiff and Vale Research and Development committees, and Cardiff University School of Psychology Research Ethics Committee. Written informed consent was obtained from all participants.

**Table 1:**
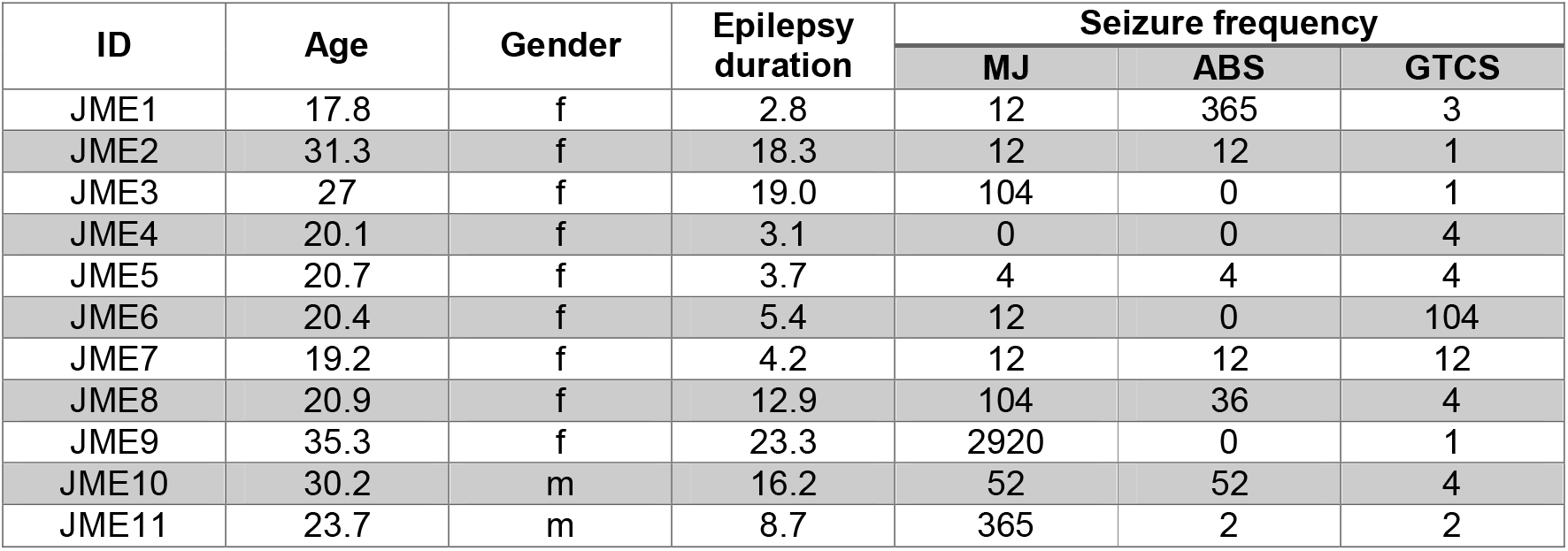
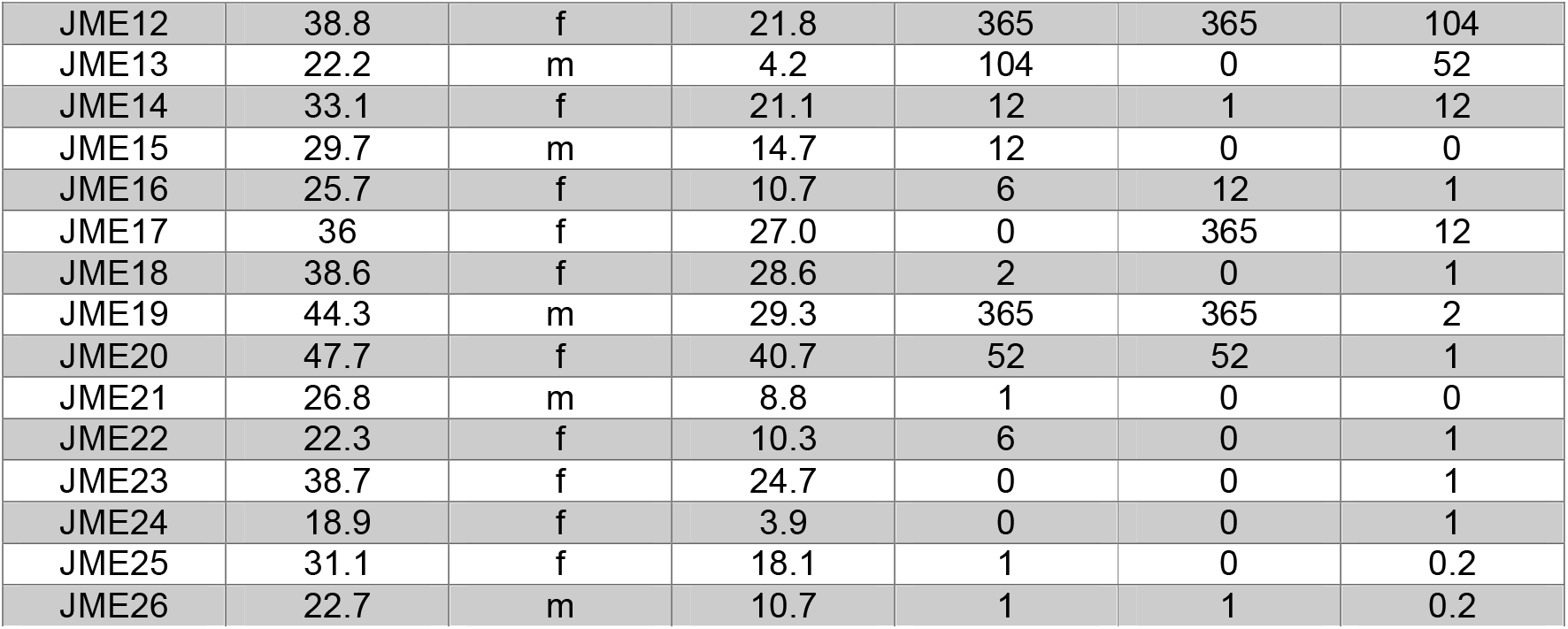
Clinical characteristics of the individuals with JME. Age and epilepsy duration are in years, m= male, f = female, seizure frequency is in number of seizures per year and is divided in three types of epileptiform activity: MJ = myoclonic jerks, ABS = absence seizures, and GTCS = generalized tonic-clonic seizures. Seizure frequency was based on self-reporting at the time of scan and extrapolated to a number of seizures per year.

| ID | Age | Gender | Epilepsy duration | Seizure frequency |  |  |
| --- | --- | --- | --- | --- | --- | --- |
|  |  |  |  | MJ | ABS | GTCS |
| JME1 | 17.8 | f | 2.8 | 12 | 365 | 3 |
| JME2 | 31.3 | f | 18.3 | 12 | 12 | 1 |
| JME3 | 27 | f | 19.0 | 104 | 0 | 1 |
| JME4 | 20.1 | f | 3.1 | 0 | 0 | 4 |
| JME5 | 20.7 | f | 3.7 | 4 | 4 | 4 |
| JME6 | 20.4 | f | 5.4 | 12 | 0 | 104 |
| JME7 | 19.2 | f | 4.2 | 12 | 12 | 12 |
| JME8 | 20.9 | f | 12.9 | 104 | 36 | 4 |
| JME9 | 35.3 | f | 23.3 | 2920 | 0 | 1 |
| JME10 | 30.2 | m | 16.2 | 52 | 52 | 4 |
| JME11 | 23.7 | m | 8.7 | 365 | 2 | 2 |
| JME12 | 38.8 | f | 21.8 | 365 | 365 | 104 |
| JME13 | 22.2 | m | 4.2 | 104 | 0 | 52 |
| JME14 | 33.1 | f | 21.1 | 12 | 1 | 12 |
| JME15 | 29.7 | m | 14.7 | 12 | 0 | 0 |
| JME16 | 25.7 | f | 10.7 | 6 | 12 | 1 |
| JME17 | 36 | f | 27.0 | 0 | 365 | 12 |
| JME18 | 38.6 | f | 28.6 | 2 | 0 | 1 |
| JME19 | 44.3 | m | 29.3 | 365 | 365 | 2 |
| JME20 | 47.7 | f | 40.7 | 52 | 52 | 1 |
| JME21 | 26.8 | m | 8.8 | 1 | 0 | 0 |
| JME22 | 22.3 | f | 10.3 | 6 | 0 | 1 |
| JME23 | 38.7 | f | 24.7 | 0 | 0 | 1 |
| JME24 | 18.9 | f | 3.9 | 0 | 0 | 1 |
| JME25 | 31.1 | f | 18.1 | 1 | 0 | 0.2 |
| JME26 | 22.7 | m | 10.7 | 1 | 1 | 0.2 |

### 2.2 MEG acquisition and pre-processing

MEG data were acquired using a 275-channel CTF radial gradiometer system (CTF System, Canada) at a sampling rate of 600 Hz. We obtained approximately 5 minutes of MEG recordings per individual. The participants were instructed to sit steadily in the MEG chair with their eyes focused on a red dot on a grey background. Each individual also underwent a whole-brain T1-weighted MRI acquired using a General Electric HDx 3T MRI scanner and an 8-channel receiver head coil (GE Healthcare, Waukesha, WI) with an axial 3D fast spoiled gradient recalled sequence (echo time 3 ms; repetition time 8 ms; inversion time 450 ms; flip angle 20º; acquisition matrix 256×192×172; voxel size 1×1×1 mm).

To assess the presence of artefacts and interictal spike wave discharges, the MEG data was divided into 2 s segments and each segment was visually inspected. Artefact-free segments were identified and re-concatenated for each individual. We thus obtained concatenated recordings with a variable length ranging from 204 s to 300 s, and to avoid the potential impact of different recording lengths on our analysis, we only considered the first 200 s of each recording for every individual. The pre-processed data were then filtered in the alpha band (8–13 Hz) and down-sampled to 250 Hz. We focused on the alpha band because it has been shown to be the most informative for differentiating people with epilepsy from healthy controls (Schmidt et al., 2014, 2016).

### 2.3 Source mapping from MEG

To infer functional networks from the MEG data, we first mapped the data from the sensor space to the source space. The MEG sensors were co-registered with the structural MRI using the locations of the fiducial coils in the CTF software (MRIViewer and MRIConverter), and we obtained a volume conduction model from the MRI scan using a semi-realistic model (Nolte, 2003). To reconstruct the source signals, we used a linear constrained minimum variance (LCMV) beamformer on a 6-mm template with a local-spheres forward model in Fieldtrip (22 Oostenveld et al., 2011; http://www.ru.nl/neuroimaging/fieldtrip). We mapped the source signals into the 90 brain regions of the Automated Anatomical Label (AAL) atlas (Hipp et al., 2012). For more details about these methods see our previous studies (Krzemiński et al., 2020, 25 Routley et al., 2020).

### 2.4 Functional networks

We divided the 200-s-long source reconstructed MEG recordings into 10, non-overlapping, 20 s segments. The choice of segment length was motivated by previous studies that aimed to distinguish people with epilepsy from controls using resting-state scalp EEG (Schmidt et al., 2014, 2016). For each segment, we computed a functional network using the amplitude envelope correlation (AEC) with orthogonalized signals (Hipp et al., 2012) (see Supplementary Material S1 for more details). We selected this method because it has been shown to be a reliable measure of functional connectivity (Colclough et al., 2016). To remove spurious connections, we generated 99 surrogates from the original MEG signals using the iterative amplitude-adjusted Fourier transform (IAAFT) with 10 iterations (Schreiber and Schmitz, 1996, 2000) (surrogates are randomized time series comparable to the original time series). We excluded connections if their weights did not exceed the 95% significance level compared to the same connection weights as computed from the surrogates (Schmidt et al., 2014, 2016, Lopes et al., 2019). Using this method, we obtained 10 functional networks per individual.

### 2.5 Mathematical model

To study the inherent propensity of a MEG functional network to generate seizures, we placed a canonical mathematical model of ictogenicity at each network node, i.e. at each of the 90 brain regions represented in the functional network (Lopes et al., 2017, 2018, 2019, 2020). The activity of a network node was described by a phase oscillator, which could transit between two states: a ‘resting state’ at which the oscillator fluctuated close to a fixed stable phase and a ‘seizure state’ represented by a rotating phase (see Supplementary Material S2 for more details about the model). This canonical model has been shown to approximate the interaction between neural masses (31 Lopes et al., 2017).

### 2.6 Brain network ictogenicity

The mathematical model allowed us to generate synthetic brain activity which fluctuated between the resting and the seizure states. To quantify this activity, we used the BNI (Chowdhury et al., 2014; Petkov et al., 2014; Lopes et al., 2017, 2018, 2019, 2020), which is the average fraction of time that the network spent in the seizure state (see Supplementary Material S3 for more details). We interpret higher values of BNI as representing a higher inherent propensity of the brain to generate seizure activity. Thus, although we use resting-state MEG data to infer the functional networks, we assume that the underlying brain states may differ in their inherent propensity to generate seizures and this may be captured by our computational framework. We expect that functional networks from JME individuals should be characterized by higher values of BNI than those from healthy individuals.

The simulated synthetic activity depends on a model parameter, the global scaling coupling (see Supplementary Material S2). Higher *K* values imply stronger neuronal interactions between connected nodes, which in turn leads to higher BNI values. Hence, for a fair comparison of BNI between different functional networks, *K* must be the same in all simulations. To avoid an arbitrary choice of *K*, we considered a redefinition of BNI (Lopes et al., 2018). This redefinition consists in computing BNI for a sufficiently large interval of *K* values in order to capture the full variation of BNI from 0 to 1. Then we calculated 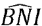 as the integral of the BNI in this interval (see Supplementary Material S3). For a meaningful comparison between different functional networks, we used the same interval of *K* for all simulations. This procedure has been shown to be robust (Lopes et al., 2018).Analogously to the BNI, a higher 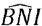 value corresponds to a higher propensity of a network to generate seizures. Figure 1 summarizes the key steps of our method.

**Figure 1.**
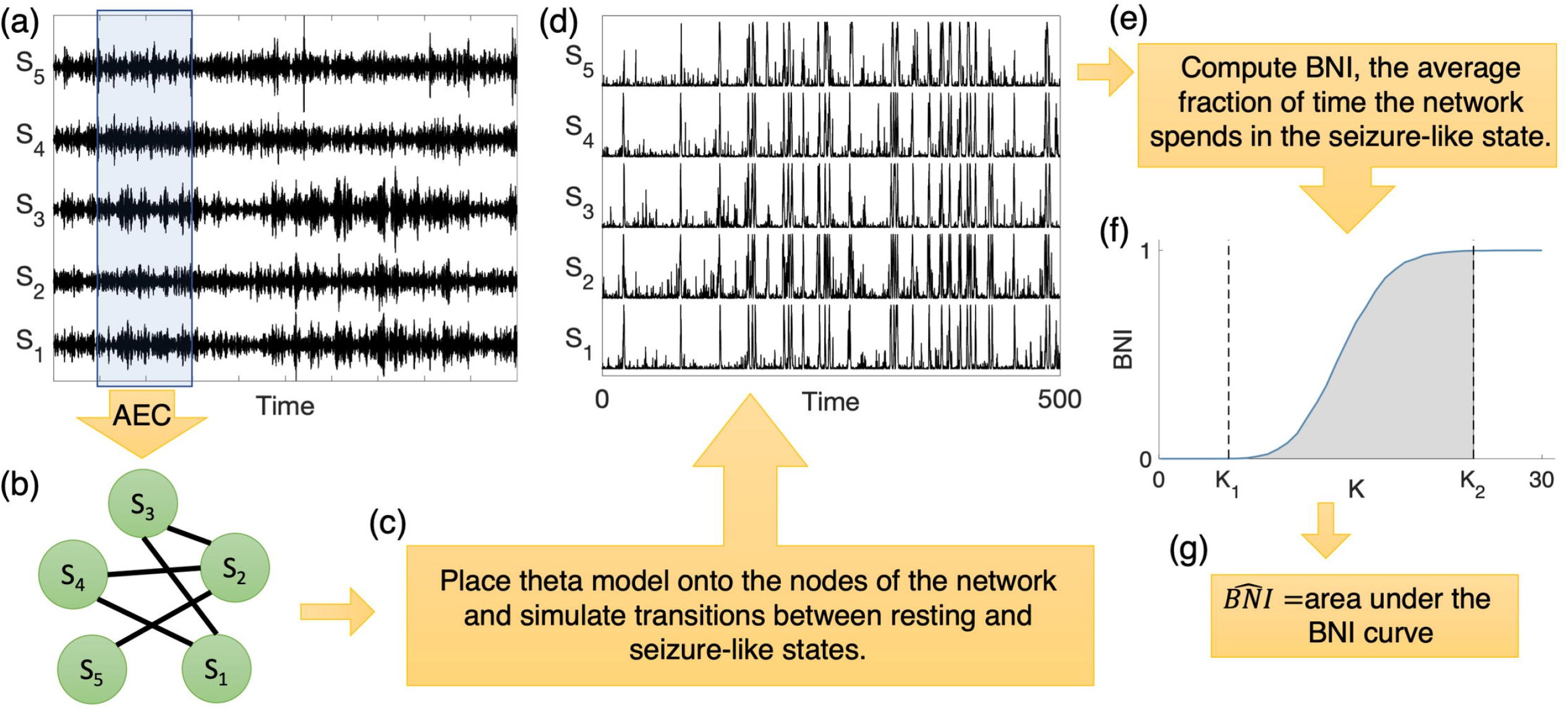
Scheme of the data analysis procedure to compute 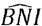. (a) We select a MEG source reconstructed data segment and by measuring the AEC we obtain (b) a functional network. To assess the propensity of the network to generate seizures, we then use (c) the theta model to simulate (d) synthetic brain activity. We then calculate (e) the BNI, i.e. the average fraction of time that network nodes spend in seizure-like activity. To avoid an arbitrary choice of *K*, we compute (f) BNI as a function of (g) 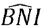 is then the integral of BNI in the interval [*K*_1_, *K*_2_] i.e. the area under the BNI curve.

### 2.7 Statistical methods

We computed 10 functional networks per individual and therefore we obtained 10 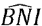 values per individual. We then calculated 〈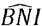〉, the average of the 10 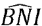 values. Finally, we used the Mann-Whitney *U* test to assess whether the median of 〈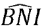〉 was higher in people with epilepsy than in the healthy controls.

## 3. Results

We considered resting-state MEG recordings from 26 people with JME and 26 healthy controls. To test whether 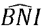 was larger in individuals with JME than in healthy controls, we first built functional networks from MEG source reconstructed data, then we placed a mathematical model of ictogenicity into the network nodes and measured the networks’ propensity to generate seizures *in silico*. Figure 2(a) shows the 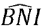 for all individuals. Overall, individuals with JME had larger 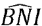 values than healthy controls (*p* = 0.0039, Mann-Whitney *U* test). This finding confirms our hypothesis that resting-state functional networks from people with epilepsy have a higher propensity to generate seizures than those from healthy controls. Note also that for each individual, we observed that 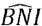 had a small variance (i.e. the intraindividual BNI variability is smaller than the interindividual BNI variability), implying that 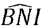 was consistent across the 10 different MEG resting-state functional networks of each individual. We then tested whether 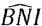 could be used for individual classification as to whether individuals had epilepsy. Figure 2(b) shows the receiver operating characteristic (ROC) curve. The area under the curve (AUC) was 0.72, the sensitivity was 0.77, and the specificity was 0.58. The 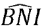’s classification accuracy was 73%.

The results in Fig. 2 may be confounded by a number of factors. Namely, epilepsy duration and seizure frequency may have an impact on the 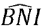 values. Figure S1 shows the 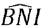 versus these clinical characteristics in the JME group. From visual inspection, the figure suggests that while individuals with short epilepsy duration or low seizure frequency may exhibit both low and high 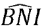 values, individuals with relatively longer epilepsy duration (larger than 20 years) and higher seizure frequency (higher than 200 seizures per year) present high 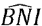 values.

**Figure 2.**
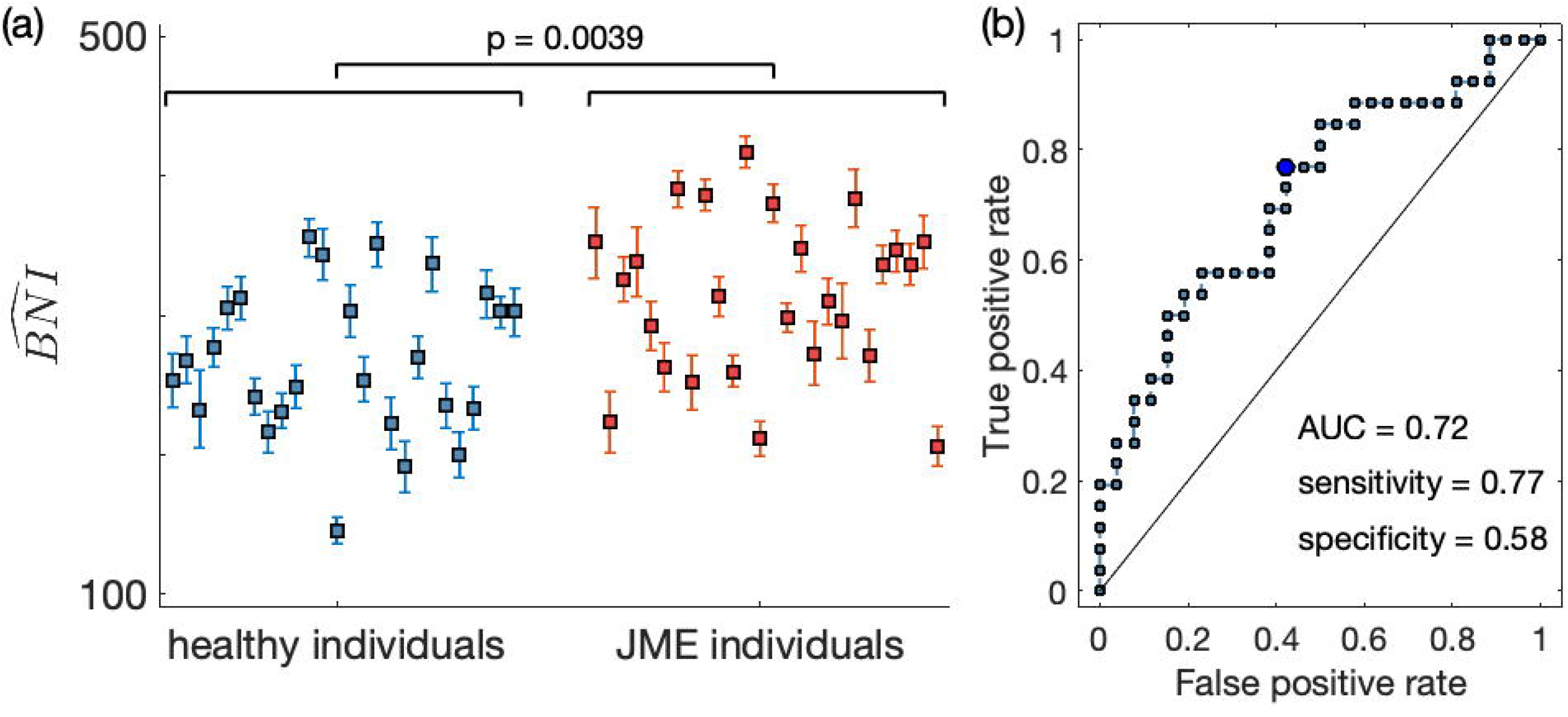
Brain network ictogenicity (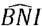) in healthy individuals and people with JME. Each marker in panel (a) represents the average 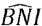 (i.e. 〈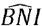〉) of a single individual and the error bars their standard error computed from 10 MEG resting-state functional networks. Blue markers correspond to healthy individuals, whereas red markers correspond to individuals with epilepsy. The epilepsy group has a larger 〈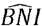〉 than the healthy group (*p* = 0.0039, Mann-Whitney *U* test). Panel (b) shows the receiver operating characteristic (ROC) curve for one group versus the other using the 〈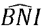〉 as a classifier. The area under the curve (AUC) is 0.72 and the circle identifies the optimal operating point of the ROC curve, for which the sensitivity is 0.77, and the specificity is 0.58.

## 4. Discussion

To date, the BNI framework has proved to be valuable for the diagnosis of IGE using scalp EEG (Schmidt et al., 2014, 2016; Petkov et al., 2014), assessment of epilepsy surgery using intracranial EEG (Goodfellow et al., 2016; Lopes et al., 2017, 2018, 2020; Laiou et al., 2019), and epilepsy classification using scalp EEG (Lopes et al., 2019). Here we aimed to test whether the concept of BNI could differentiate people with JME from age and gender matched healthy controls using resting-state MEG data. We found that the BNI is on average higher in the JME group than in the control group.We further found that, as a classifier, the BNI yields a sensitivity of 0.77, a specificity of 0.58, and an AUC of 0.72, which is similar to previous findings in IGE using scalp EEG (Schmidt et al., 2016). This result suggests that MEG and scalp EEG may yield similar diagnostic power despite MEG being considered superior to EEG in recording reliable brain signals (Supek and Aine, 2016). In other words, this suggests that the key functional network properties that characterize the underlying brain ictogenicity may be similarly captured by MEG and scalp EEG.

Resting-state MEG functional networks have been previously shown to be altered in people with epilepsy relative to healthy controls (van Dellen et al., 2012; Niso et al., 2015; Hsiao et al., 2015; Wu et al., 2017; Routley et al., 2020). For example, Niso et al. (2015) used 15 graph-theoretic measures to quantify resting-state MEG functional networks from people with frontal focal epilepsy, generalized epilepsy and healthy individuals. They found that functional networks from generalized epilepsy had greater efficiency and lower eccentricity than those from controls, whereas functional networks from frontal focal epilepsy exhibited only reduced eccentricity over fronto-temporal and central sensors relative to networks from controls. Furthermore, machine learning has been used to also differentiate people with epilepsy from controls (Soriano et al., 2017). Our study distinguishes from these studies by not only searching for differences between functional networks in health and disease, but instead test a specific mechanistic hypothesis that justifies the difference. Thus, our approach is more readily interpretable and may offer insight into why altered functional networks underlie epilepsy.

We acknowledge that our study has some limitations. First, in order to truly test how MEG-based predictions compare to scalp EEG-based predictions, we would need both MEG and EEG data collected from the same participants. Future work should assess whether predictions based on both data modalities would deliver equivalent individual classification. Second, people with JME were taking anti-epileptic medication, which may have potentially reduced the BNI in some JME individuals, making them indistinguishable from healthy individuals. Future studies should consider newly diagnosed drug-naïve individuals. This may be particularly important to also control for the effect of epilepsy duration and seizure frequency on BNI. Our results suggest that individuals with longer epilepsy duration and higher seizure frequency were more likely to be characterized by high BNI. On one hand this is an expected observation, i.e. BNI should be higher for individuals more prone to seizures and also those for which a longer disease may have had an impact on resting-state functional connectivity. On the other hand, these were individuals for which diagnosis could be less challenging. Third, we focused our analysis on differentiation of people with JME from healthy controls. We therefore cannot exclude the possibility that our findings are specific to JME. More comprehensive datasets will be needed to explore whether our findings generalize to other types of epilepsy.

## 5. Conclusions

Our results demonstrate that the BNI framework generalizes from scalp EEG to MEG. We showed that resting-state MEG from people with JME is characterized by higher BNI than that from healthy controls. The achieved classification accuracy is similar to previously obtained from scalp EEG, suggesting that the two data modalities may capture similar underlying ictogenic features.

## Data Availability

All materials (functional networks) are available upon request (contact).

## Acknowledgements

ML gratefully acknowledges funding from Cardiff University’s Wellcome Trust Institutional Strategic Support Fund (ISSF) [204824/Z/16/Z]. DK was supported by an EPSRC PhD studentship [grant number EP/N509449/1]. KH acknowledges support from the Health and Care Research Wales:Clinical Research Time Award and the Wales BRAIN Unit. JT acknowledges the financial support of the EPSRC via grant EP/N014391/2 and a Wellcome Trust Institutional Strategic Support Award(WT105618MA). JZ acknowledges the financial support of the European Research Council [grantnumber 716321]. KDS and KH acknowledge the support of the UK MEGMRC Partnership Grant(MRC/EPSRC, MR/K005464/1) and a Wellcome Trust Strategic Award (104943/Z/14/Z).

## References

1 Abela, E.,Pawley, A. D., Tangwiriyasakul, C., Yaakub, S. N., Chowdhury,F. A., Elwes, R. D., … & Richardson, M. P.(2019). Slower alpha rhythm associates with poorer seizure control in epilepsy. Annals of clinical and translational neurology, 6(2), 333–343. doi: 10.1002/acn3.710

2 Benbadis, S.(2009). The differential diagnosis of epilepsy: a critical review. Epilepsy & Behavior, 15(1), 15–21. doi: 10.1016/j.yebeh.2009.02.024

3 Benbadis, S. R., Beniczky, S., Bertram, E.,MacIver, S., &Moshé, S. L.(2020). The role of EEG in patients with suspected epilepsy. Epileptic Disorders, 22(2), 143–155. doi: 10.1684/epd.2020.1151

4 Chowdhury, F. A., Woldman, W., FitzGerald, T. H.,Elwes, R. D., Nashef, L.,Terry, J. R.,& Richardson, M. P. (2014). Revealing a brain network endophenotype in families with idiopathic generalised epilepsy. PloS one, 9(10), e110136. doi: 10.1371/journal.pone.0110136

5 Colclough, G. L., Woolrich, M. W., Tewarie, P. K., Brookes, M. J., Quinn, A. J., & Smith, S. M. (2016). How reliable are MEG resting-state connectivity metrics?. Neuroimage, 138, 284–293. doi: 10.1016/j.neuroimage.2016.05.070

6 Coito, A., Genetti, M., Pittau, F., Iannotti, G. R., Thomschewski, A.,Höller, Y., … & Plomp, G., (2016). Altered directed functional connectivity in temporal lobe epilepsy in the absence of interictal spikes: a high density EEG study. Epilepsia, 57(3), 402–411. doi: 10.1111/epi.13308

7 van Dellen, E., Douw, L., Hillebrand, A., Ris-Hilgersom, I. H., Schoonheim, M. M., Baayen, J. C., … & Stam, C. J., (2012). MEG network differences between low-and high-grade glioma related to epilepsy and cognition. PloS one, 7(11). doi: 10.1371/journal.pone.0050122

8 Hipp, J. F., Hawellek, D. J., Corbetta, M., Siegel, M., & Engel, A. K., (2012). Large-scale cortical correlation structure of spontaneous oscillatory activity. Nature neuroscience, 15(6), 884. doi: 10.1038/nn.3101

9 Horstmann, M. T., Bialonski, S., Noennig, N., Mai, H., Prusseit, J., Wellmer, J., … & Lehnertz, K. (2010). State dependent properties of epileptic brain networks: Comparative graph–theoretical analyses of simultaneously recorded EEG and MEG. Clinical Neurophysiology, 121(2), 172–185. doi: 10.1016/j.clinph.2009.10.013

10 Hsiao, F. J., Yu, H. Y., Chen, W. T., Kwan, S. Y., Chen, C., Yen, D. J., … & Lin, Y. Y. (2015).Increased intrinsic connectivity of the default mode network in temporal lobe epilepsy: evidence from resting-state MEG recordings. PLoS One, 10(6). doi: 10.1371/journal.pone.0128787

11 Krzemiński, D., Masuda, N., Hamandi, K., Singh, K. D., Routley, B., & Zhang, J., (2020). Energy landscape of resting magnetoencephalography reveals fronto-parietal network impairments in epilepsy. Network Neuroscience, 1–23. doi: 10.1162/netn_a_00125

12 Laiou, P., Avramidis, E., Lopes, M. A., Abela, E.,Müller, M., Akman, O. E., … & Goodfellow, M., (2019). Quantification and selection of ictogenic zones in epilepsy surgery. Frontiers in neurology, 10,1045. doi: 10.3389/fneur.2019.01045

13 Larsson, P. G., & Kostov, H., (2005). Lower frequency variability in the alpha activity in EEG among patients with epilepsy. Clinical Neurophysiology, 116(11), 2701–2706. doi: 10.1016/j.clinph.2005.07.019

14 Lopes, M. A., Richardson, M. P., Abela, E., Rummel, C., Schindler, K., Goodfellow, M., & Terry, J. R., (2017). An optimal strategy for epilepsy surgery: Disruption of the rich-club?. PLoS computational biology, 13(8), e1005637.doi: 10.1371/journal.pcbi.1005637

15 Lopes, M. A., Richardson, M. P., Abela, E., Rummel, C., Schindler, K., Goodfellow, M., & Terry, J. R., (2018). Elevated ictal brain network ictogenicity enables prediction of optimal seizure control.Frontiers in neurology, 9, 98. doi: 10.3389/fneur.2018.00098

16 Lopes, M. A., Perani, S., Yaakub, S. N., Richardson, M. P., Goodfellow, M., & Terry, J. R., (2019). Revealing epilepsy type using a computational analysis of interictal EEG. Scientific reports, 9(1), 1–10. doi: 10.1038/s41598-019-46633-7

17 Lopes, M. A., Junges, L., Tait, L., Terry, J. R., Abela, E., Richardson, M. P., & Goodfellow, M., (2020). Computational modelling in source space from scalp EEG to inform presurgical evaluation of epilepsy. Clinical Neurophysiology, 131(1), 225–234. doi: 10.1016/j.clinph.2019.10.027

18 Niso, G., Carrasco, S., Gudín, M., Maestú, F., del-Pozo, F., & Pereda, E., (2015). What graph theory actually tells us about resting state interictal MEG epileptic activity. Neuroimage: clinical, 8, 503–515. doi: 10.1016/j.nicl.2015.05.008

19 Nolte, G. (2003). The magnetic lead field theorem in the quasi-static approximation and its use for magnetoencephalography forward calculation in realistic volume conductors. Physics in Medicine & Biology, 48(22),3637. doi: 10.1088/0031-9155/48/22/002

20 Oostenveld, R., Fries, P., Maris, E., & Schoffelen, J. M., (2011). FieldTrip: open source software for advanced analysis of MEG,EEG, and invasive electrophysiological data. Computational intelligence and neuroscience,2011, 1. doi: 10.1155/2011/156869

21 Petkov, G., Goodfellow, M., Richardson, M. P., & Terry, J. R. (2014). A critical role for network structure in seizure onset: a computational modeling approach. Frontiers in neurology, 5, 261. doi: 10.3389/fneur.2014.00261

22 Pillai, J., & Sperling, M. R., (2006). Interictal EEG and the diagnosis of epilepsy. Epilepsia, 47, 14–22. doi: 10.1111/j.1528-1167.2006.00654.x

23 Quraan, M. A., McCormick, C., Cohn, M., Valiante, T. A., & McAndrews, M. P. (2013). Altered resting state brain dynamics in temporal lobe epilepsy can be observed in spectral power, functional connectivity and graph theory metrics. PloS one, 8(7). doi: 10.1371/journal.pone.0068609

24 Routley, B., Shaw, A., Muthukumaraswamy, S. D., Singh, K. D., & Hamandi, K., (2020). Juvenile myoclonic epilepsy shows increased posterior theta, and reduced sensorimotor beta resting connectivity. Epilepsy Research, 106324. doi: 10.1016/j.eplepsyres.2020.106324

25 Schmidt H., Petkov G.,., Richardson M.P., Terry J.R. (2014) Dynamics on networks: the role of local dynamics and global networks on the emergence of hypersynchronous neural activity. PLoS Comput Biol, 10:e1003947. doi: 10.1371/journal.pcbi.1003947

26 Schmidt H., Woldman W., Goodfellow M., Chowdhury F.A., Koutroumanidis M., Jewell S., et al. (2016) A computational biomarker of idiopathic generalized epilepsy from resting state EEG. Epilepsia, 57:e200–4. doi: 10.1111/epi.13481

27 Schreiber, T., & Schmitz, A., (1996). Improved surrogate data for nonlinearity tests. Physical review letters, 77(4), 635. doi: 10.1103/PhysRevLett.77.635

28 Schreiber, T., & Schmitz, A., (2000). Surrogate time series. Physica D: Nonlinear Phenomena, 142(3–4), 346–382.doi: 10.1016/S0167-2789(00)0004-9

29 Smith, S. J. M., (2005). EEG in the diagnosis, classification, and management of patients with epilepsy. Journal of Neurology, Neurosurgery & Psychiatry, 76(suppl 2), ii2–ii7. doi: 10.1136/jnnp.2005.069245

30 Soriano, M. C., Niso, G., Clements, J., Ortín, S., Carrasco, S., Gudín, M., … & Pereda, E., (2017). Automated detection of epileptic biomarkers in resting-state interictal MEG data. Frontiers in neuroinformatics, 11, 43. doi: 10.3389/fninf.2017.00043

31 Supek, S., & Aine, C. J., (2016). Magnetoencephalography. Springer-Verlag Berlin An. doi: 10.1007/978-3-642-33045-2

32 van Diessen, E., Otte, W. M., Braun, K. P., Stam, C. J., & Jansen, F. E., (2013). Improved diagnosis in children with partial epilepsy using a multivariable prediction model based on EEG network characteristics. PloS one, 8(4). doi: 10.1371/journal.pone.0059764

33 van Diessen, E., Otte, W. M., Stam, C. J., Braun, K. P., & Jansen, F. E., (2016). Electroencephalography based functional networks in newly diagnosed childhood epilepsies. Clinical Neurophysiology, 127(6), 2325–2332. doi: 10.1016/j.clinph.2016.03.015

34 Verhoeven, T., Coito, A., Plomp, G., Thomschewski, A., Pittau, F., Trinka, E., … & Dambre, J., (2018). Automated diagnosis of temporal lobe epilepsy in the absence of interictal spikes. NeuroImage: Clinical,17, 10–15. doi: 10.1016/j.nicl.2017.09.021

35 WHO(2019)http://www.who.int/news-room/fact-sheets/detail/epilepsy

36 Wu, C., Xiang, J., Jiang, W., Huang, S., Gao, Y., Tang, L., … & Wang, X., (2017). Altered effective connectivity network in childhood absence epilepsy: a multi-frequency MEG study. Brain topography, 30(5), 673–684. doi: 10.1007/s10548-017-0555-1

